# Risk of Neuroendocrine Neoplasms with Glucagon-Like-Petide-1 Receptor Agonist Use in Patients with Type 2 Diabetes Mellitus

**DOI:** 10.1101/2025.07.18.25331594

**Authors:** Gordon Hong, Dipen Patel, Abbinaya Elangovan, Bhavana Konda, Raj Shah, Vineeth Sukrithan

**Affiliations:** Department of Medicine, Case Western Reserve University/University Hospitals Cleveland Medical Center, Cleveland, Ohio, USA; Division of Medical Oncology, Department of Internal Medicine, The Ohio State University Comprehensive Cancer Center, Columbus, Ohio, USA; Division of Gastroenterology, Department of Medicine, Virginia Tech Carilion Clinic, Roanoke, Virginia, USA; Division of Gastroenterology, Hepatology, and Nutrition, Department of Internal Medicine, The Ohio State University Wexner Medical Center, Columbus, OH, USA

**Author notes:** Denotes co-last author (Raj Shah, Vineeth Sukrithan). **Corresponding Author:** Vineeth Sukrithan, MD, Suite 1335 Lincoln Tower, 1800 Cannon Dr., Columbus, OH 43210.

## Abstract

**Introduction:** Prior studies demonstrate reduced risk of several malignancies among patients with type 2 diabetes mellitus (T2DM) prescribed glucagon-like-peptide-1 receptor agonists (GLP-1RA). A recent study however found expression of GLP-1R among neuroendocrine neoplasm (NEN) cell lines and growth promotion in xenografts treated with semaglutide. In this study, we evaluate incidence of NENs among patients diagnosed as overweight/obese and patients with T2DM prescribed GLP-1RA based drugs.

**Methods:** The TriNetX database was utilized for this study. Incidence of NENs between patients with overweight/obesity were compared to a control group without obesity within 15 years following index diagnosis of obesity or, in the control group, a non-specific outpatient visit. Groups were matched for demographics, tobacco use, family malignancy history, alcohol use, and socioeconomic factors. In a separate analysis, incidence of NENs among patients with T2DM were compared between those prescribed GLP-1RAs versus other antihyperglycemic agents (AHA) within 15 years following index prescription of GLP-1RA or comparator AHA. Patients with exposure to the comparator AHA were excluded from their respective groups. A subgroup analysis was performed among patients with overweight/obesity. Exclusion criteria included Merkel cell carcinoma, multiple endocrine neoplasia type 1, Cushing’s syndrome, Von Hippel-Lindau syndrome, tuberous sclerosis, and prior NEN.

**Results:** Patients with overweight/obesity had a significant increased risk of NENs when compared to the control group (3,840/794,235 (0.48%) vs. 2,785/794,235 (0.35%); RR: 1.38 (95% CI 1.31-1.45), *P* < 0.0001). Among patients with T2DM, GLP-1RA exposure was associated with significantly lower risk of NENs compared to insulin (70/31,171 (0.23%) vs. 137/31,171 (0.44%); RR: 0.51 (0.38, 0.68), *P* < 0.0001), but no significant difference was appreciated with other AHAs. In the subset of patients with obesity/overweight, GLP-1RAs were associated with decreased risk compared to insulin (43/15,230 (0.28%) vs. 68/15,230 (0.45%); RR: 0.63 (0.43, 0.93), p=0.02) and metformin (39/10,370 (0.38%) vs. 64/10,370 (0.62%); RR: 0.61 (0.41, 0.91), *P* = 0.01).

**Conclusion:** This study suggests obesity to be significantly associated with incidence of NENs. Furthermore, our findings do not demonstrate an increased incidence of NENs with GLP-RA use among patients with T2DM and show decreased risk when compared to insulin exposure.

## Introduction

Recent studies demonstrate significant risk reduction in obesity-related cancers among patients with type 2 diabetes mellitus (T2DM) prescribed glucagon-like-peptide-1 receptor agonists (GLP-1RA) compared to insulin.^1^ T2DM and obesity are known risk factors for neuroendocrine neoplasms (NENs).^2^ Specifically, visceral obesity, dyslipidemia, and fasting hyperglycemia are associated with up to 4 times greater risk of gastroenteropancreatic-NENs (GEP-NENs).^3^ Medications treating obesity such as GLP-1RAs may potentially improve metabolic parameters that drive tumor growth. However, one study indicated that 50% of NEN cell lines expressed GLP-1R, and growth promotion was observed in xenografts of GEP-NENs treated with semaglutide, a GLP-1RA.^4^ Given that randomized trials of GLP-1RAs on NENs are unlikely to be forthcoming, we sought to study the incidence of NENs with GLP-1RA-based drugs using TriNetX, a global, real-world electronic health record database.

## Methods

We compared the incidence of NENs between patients diagnosed with being overweight or obese and a control group of patients without a diagnosis of overweight/obese. Groups were propensity-score matched for demographics, tobacco use, family malignancy history, alcohol use, and socioeconomic factors. Patients were evaluated for diagnosis of NENs within 15 years following index diagnosis of overweight/obesity or, in the control group, index event of a non-specific outpatient visit. Patients were required to have an outpatient visit and general health screening within the follow-up period to be included. Subsequently, we evaluated the incidence of NENs in patients with T2DM prescribed GLP-1RAs versus other antihyperglycemic agents (AHA) with matching for the same factors described previously with the addition of overweight/obesity **(Table 1)**. Patients in the GLP-1RA group with exposure to the comparison drug were excluded. The index event in each analysis was prescription of GLP-1RA or comparison medication. A subgroup analysis was also performed within the overweight/obese population. Exclusion criteria included Merkel cell carcinoma, multiple endocrine neoplasia type 1, Cushing’s syndrome, Von Hippel-Lindau syndrome, tuberous sclerosis, and prior NEN. Analyses were performed using the built-in statistics program of TriNetX to identify incidence rates and relative risks (RR). Since patient data on this platform is deidentified, this study is exempt from institutional review board approval.

**Table 1:** Cohort Characteristics Prior to Propensity Score Matching. Demographic and characteristic data of cohorts included in the analysis of incidence of neuroendocrine neoplasms between patients with exposure to glucagon-like-peptide-1 receptor agonists (GLP-1RA) compared to other antihyperglycemic agents prior to propensity score matching. DPP4i indicate dipeptidyl-peptidase 4 inhibitors, SGLT2i: sodium-glucose cotransporter 2 inhibitors, SU: sulfonylureas, TZD: thiazolidinediones. (+) indicates exposure to medication is present and (-) indicates exposure to medication is excluded in the cohort.

|  | GLP1-RA(+)/insulin(-)<br>(n=31,174) | GLP1-RA(-)/insulin(+)<br>(n=1,131,709) | GLP1-RA(+)/metformin(-)<br>(n=20,854) | GLP1-RA(-)/metformin(+)<br>(n=1,021,969) | GLP1-RA(+)/DPP4i(-)<br>(n=64,838) | GLP1-RA(-)/DPP4i(+)<br>(n=140,822) |
| --- | --- | --- | --- | --- | --- | --- |
| Age at Index Event, mean (SD), years | 55.2 ± 11.9 | 59.2 ± 14.1 | 58.7 ± 12.2 | 59.2 ± 12.7 | 56.8 ± 11.8 | 63.1 ± 11.4 |
| Female Sex | 17,619 (56.52%) | 507,075 (44.98%) | 11,810 (56.64%) | 484,428 (47.71%) | 34,539 (53.27%) | 64,389 (45.72%) |
| Ethnicity |  |  |  |  |  |  |
| Hispanic or Latino | 4,059 (13.02%) | 106,131 (9.42%) | 1,784 (8.56%) | 109,012 (10.74%) | 6,709 (10.35%) | 11,268 (8.00%) |
| Not Hispanic or Latino | 20,995 (67.35%) | 736,582 (65.34%) | 14,018 (67.22%) | 666,469 (65.64%) | 44,182 (68.14%) | 94,784 (67.31%) |
| Unknown Ethnicity | 6,118 (19.63%) | 284,543 (25.24%) | 5,051 (24.22%) | 239,825 (23.62%) | 13,947 (21.51%) | 34,770 (24.69%) |
| Race |  |  |  |  |  |  |
| White | 21,182 (67.95%) | 656,213 (58.21%) | 13,706 (65.73%) | 593,976 (58.50%) | 44,643 (68.85%) | 90,411 (57.10%) |
| Black/African American | 4,984 (15.99%) | 227,766 (20.21%) | 3,448 (16.54%) | 176,707 (17.40%) | 9,693 (14.95%) | 20,588 (14.62%) |
| Asian | 764 (2.45%) | 64,004 (5.68%) | 431 (2.07%) | 63,567 (6.26%) | 1,398 (2.16%) | 12,777 (9.07%) |
| Other Race | 896 (2.87%) | 43,739 (3.88%) | 753 (3.61%) | 46,035 (4.53%) | 2,040 (3.15%) | 6,196 (4.40%) |
| Native Hawaiian or Pacific Islander | 88 (0.28%) | 12,174 (1.08%) | 86 (0.41%) | 4,472 (0.44%) | 206 (0.32%) | 553 (0.39%) |
| American Indian or Alaska Native | 157 (0.50%) | 3,906 (0.35%) | 76 (0.36%) | 3,795 (0.37%) | 283 (0.44%) | 427 (0.30%) |
| Tobacco use | 621 (1.99%) | 15,985 (1.42%) | 252 (1.21%) | 12,914 (1.27%) | 955 (1.47%) | 959 (0.68%) |
| Family History of Malignancy | 681 (2.19%) | 19,797 (1.76%) | 523 (2.51%) | 19,383 (1.91%) | 1,304 (2.01%) | 1,689 (1.20%) |
| Overweight and Obesity | 6,364 (20.42%) | 126,914 (11.26%) | 4,192 (20.10%) | 120,045 (11.82%) | 11,626 (17.93%) | 9,971 (7.08%) |
| Alcohol related disorders | 209 (0.67%) | 29,643 (2.63%) | 142 (0.68%) | 17,896 (1.76%) | 376 (0.58%) | 1,015 (0.72%) |
| Adverse socioeconomic determinants of health | 22 (0.07%) | 4,612 (0.41%) | 16 (0.08%) | 2,732 (0.27%) | 52 (0.08%) | 95 (0.07%) |

**Table 1:** Demographic and characteristic data of cohorts included in the analysis of incidence of neuroendocrine neoplasms between patients with exposure to glucagon-like-peptide-1 receptor agonists (GLP-1RA) compared to other antihyperglycemic agents prior to propensity score matching.
|  | GLP1-RA(+)/SGLT2i(-)<br>(n=47,739) | GLP1-RA(-)/SGLT2i(+)<br>(n=26,285) | GLP1-RA(+)/SU(-)<br>(n=48,565) | GLP1-RA(-)/SU(+)<br>(n=389,756) | GLP1-RA(+)/TZD(-)<br>(n=64,432) | GLP1-RA(-)/TZD(+)<br>(n=71,839) |
| --- | --- | --- | --- | --- | --- | --- |
| Age at Index Event, mean (SD), years | 57.2 ± 12.1 | 59.3 ± 11.3 | 56.5 ± 11.9 | 61.17 ± 12.1 | 56.9 ± 11.8 | 61.7 ± 11 |
| Female Sex | 26,591 (55.70%) | 10,183 (38.74%) | 26,713 (55.01%) | 170,997 (44.59%) | 35,101 (54.48%) | 30,046 (42.49%) |
| Ethnicity |  |  |  |  |  |  |
| Hispanic or Latino | 5,179 (10.85%) | 2,233 (8.50%) | 4,866 (10.02%) | 42,756 (11.15%) | 6,212 (9.64%) | 8,088 (11.44%) |
| Not Hispanic or Latino | 33,552 (70.28%) | 17,364 (66.06%) | 32,709 (67.35%) | 251,501 (65.58%) | 43,711 (67.84%) | 43,740 (67.51%) |
| Unknown Ethnicity | 9,008 (18.87%) | 6,688 (25.44%) | 10,988 (22.63%) | 89,238 (23.27%) | 14,507 (22.52%) | 14,855 (21.05%) |
| Race |  |  |  |  |  |  |
| White | 32,883 (68.88%) | 16,894 (64.27%) | 32,605 (67.14%) | 224,992 (58.67%) | 43,720 (67.86%) | 43,428 (61.41%) |
| Black/African American | 7,811 (16.36%) | 2,645 (10.06%) | 7,503 (15.45%) | 68,746 (17.83%) | 10,009 (15.54%) | 10,586 (14.97%) |
| Asian | 1,039 (2.18%) | 1,713 (6.52%) | 1,104 (2.27%) | 25,736 (6.71%) | 1,348 (2.09%) | 4,527 (6.40%) |
| Other Race | 1,476 (3.09%) | 1,235 (4.70%) | 1,724 (3.55%) | 15,701 (4.09%) | 2,114 (3.28%) | 2,715 (3.84%) |
| Native Hawaiian or Pacific Islander | 138 (0.29%) | 82 (0.31%) | 162 (0.33%) | 1,767 (0.46%) | 208 (0.32%) | 303 (0.43%) |
| American Indian or Alaska Native | 219 (0.46%) | 75 (0.29%) | 208 (0.43%) | 1,320 (0.34%) | 278 (0.43%) | 289 (0.41%) |
| Tobacco use | 883 (1.85%) | 256 (0.97%) | 684 (1.41%) | 2,865 (0.75%) | 900 (1.40%) | 328 (0.46%) |
| Family History of Malignancy | 1,133 (2.37%) | 398 (1.51%) | 1,111 (2.29%) | 4,243 (1.11%) | 1,274 (1.98%) | 704 (0.10%) |
| Overweight and Obesity | 9,766 (20.46%) | 2,383 (9.07%) | 9,185 (18.91%) | 29,433 (7.68%) | 11,200 (17.383%) | 5,274 (7.46%) |
| Alcohol related disorders | 319 (0.67%) | 208 (0.79%) | 296 (0.61%) | 2,865 (0.75%) | 354 (0.55%) | 545 (0.77%) |
| Adverse socioeconomic determinants of health | 48 (0.10%) | 13 (0.05%) | 42 (0.09%) | 580 (0.15%) | 43 (0.07%) | 54 (0.08%) |

## Results

Patients with overweight/obesity diagnoses had a statistically significant increased risk of NENs when compared to patients without these diagnoses (3,840/794,235 (0.48%) vs. 2,785/794,235 (0.35%); RR: 1.38 (95% CI 1.31-1.45), *P* < 0.0001). Among patients with T2DM, GLP-1RA exposure was associated with significantly lower risk of NENs compared to insulin (70/31,171 (0.23%) vs. 137/31,171 (0.44%); RR: 0.51 (0.38, 0.68), *P* < 0.0001), but no significant difference was observed when compared to metformin, dipeptidyl peptidase-4 inhibitors, sodium-glucose cotransporter 2 inhibitors, sulfonylureas, or thiazolidinediones. In a subset of patients with T2DM and obesity/overweight, exposure to GLP-1RAs was associated with decreased risk of NENs compared to both insulin (43/15,230 (0.28%) vs. 68/15,230 (0.45%); RR: 0.63 (0.43, 0.93), *P* = 0.02) and metformin (39/10,370 (0.38%) vs. 64/10,370 (0.62%); RR: 0.61 (0.41, 0.91), *P* = 0.01) **(Table 2)**.

**Table 2:** Incidence of Neuroendocrine Neoplasms in T2DM Patients with GLP-1RA Use Compared to Other Antidiabetic Medications. Comparison of incidence of NENs among patients with T2DM that were previously antidiabetic drug-naïve within 15 years of first exposure to GLP-1RAs versus other antihyperglycemic agents. Patients were matched for demographic data (age at index event, gender, race, ethnicity), overweight/obesity, tobacco use, family malignancy history, alcohol use, and socioeconomic factors. Patients with GLP-1RA exposure were at statistically significant decreased relative risk for incidence of NENs when compared to insulin in the overall study population and overweight/obesity population and when compared to metformin in the overweight/obesity population (bolded).

|  |  |  |  |  | **A:**<br>**Overall**<br>**Study**<br>**Population** |
| --- | --- | --- | --- | --- | --- |
| **Exposure cohort** | **Comparison cohort** | <u>Exposure cohort</u><br>**No. of cases (overall risk)** | <u>Comparison cohort</u><br>**No. of cases (overall risk)** | **RR (95% CI)** |  |
| **GLP1-RA(+)/insulin(-)** | **GLP1-RA(-)/insulin(+)** | **70/31,171 (0.23%)** | **137/31,171 (0.44%)** | **0.51 (0.38, 0.68)** |  |
| GLP1-RA(+)/metformin(-) | GLP1-RA(-)/metformin(+) | 71/20,853 (0.34%) | 85/20,853 (0.41%) | 0.84 (0.61, 1.14) |  |
| GLP1-RA(+)/DPP4i(-) | GLP1-RA(-)/DPP4(+) | 267/60,469 (0.44%) | 260/60,469 (0.43%) | 1.03 (0.87, 1.22) |  |
| GLP1-RA(+)/SGLT2i(-) | GLP1-RA(-)/SGLT2i(+) | 102/25,198 (0.41%) | 103/25,198 (0.41%) | 0.99 (0.75, 1.30) |  |
| GLP1-RA(+)/SU(-) | GLP1-RA(-)/SU(+) | 197/48,563 (0.41%) | 218/48,563 (0.45%) | 0.90 (0.75, 1.10) |  |
| GLP1-RA(+)/TZD(-) | GLP1-RA(-)/TZD(+) | 228/50,633 (0.45%) | 231/50,633 (0.46%) | 0.99 (0.82, 1.19) |  |

**Table 2:** Comparison of incidence of NENs among patients with T2DM that were previously antidiabetic drug-naïve within 15 years of first exposure to GLP-1RAs versus other antihyperglycemic agents.
| **Exposure cohort** | **Comparison cohort** | <u>Exposure cohort</u><br>**No. of cases (overall risk)** | <u>Comparison cohort</u><br>**No. of cases (overall risk)** | **RR (95% CI)** |
| --- | --- | --- | --- | --- |
| **GLP1-RA(+)/insulin(-)** | **GLP1-RA(-)/insulin(+)** | **43/15,230 (0.28%)** | **68/15,230 (0.45%)** | **0.63 (0.43, 0.93)** |
| **GLP1-RA(+)/metformin(-)** | **GLP1-RA(-)/metformin(+)** | **39/10,370 (0.38%)** | **64/10,370 (0.62%)** | **0.61 (0.41, 0.91)** |
| GLP1-RA(+)/DPP4i(-) | GLP1-RA(-)/DPP4(+) | 139/25,645 (0.54%) | 131/25,645 (0.51%) | 1.06 (0.84, 1.35) |
| GLP1-RA(+)/SGLT2i(-) | GLP1-RA(-)/SGLT2i(+) | 35/7,357 (0.48%) | 35/7,357 (0.48%) | 1.00 (0.63, 1.60) |
| GLP1-RA(+)/SU(-) | GLP1-RA(-)/SU(+) | 108/23,481 (0.46%) | 131/23,481 (0.56%) | 0.82 (0.64, 1.06) |
| GLP1-RA(+)/TZD(-) | GLP1-RA(-)/TZD(+) | 112/19,145 (0.59%) | 116/19,145 (0.61%) | 0.97 (0.75, 1.25) |

## Discussion

This large database study confirms obesity to be a significant predictor for incidence of NENs over a 15-year period. Reassuringly, among patients with T2DM treated with GLP-1RAs, a reduced incidence of NENs was noted compared to those on insulin and no difference was appreciated when compared to other AHAs. Additionally, in the overweight/obesity subanalysis, the incidence of NENs was reduced when compared to both insulin and metformin.

GLP-1RAs have complex pleiotropic effects on the tumor-immune microenvironment. These agents have direct effects on immune cell populations in addition to anti-inflammatory effects.^5,6^ Therefore, caution must be exercised when extrapolating results from xenograft studies in immune-deficient mice to the general population. Limitations of our study include biases inherent to retrospective studies including unmeasured confounders, selection bias, and reverse causality. External independent validation is warranted to confirm these findings.

## Data Availability

Data used for this manuscript are from the TrinetX Database available from https://trinetx.com/.

## Data Sharing Statement

Data used for this manuscript are from the TrinetX Database available from https://trinetx.com/.

## Author Contributions

V.S. : Conceptualization, writing, supervision, data collection, data interpretation

R.S. : Conceptualization, writing, supervision, data collection, data interpretation

B.K. : Supervision, Editing, data interpretation D.P: Editing, Supervision, data interpretation

G.H.: Data Collection, Data Analysis, Writing

A.E. : Data Collection, Editing

---

**Supplemental Figure 1:**
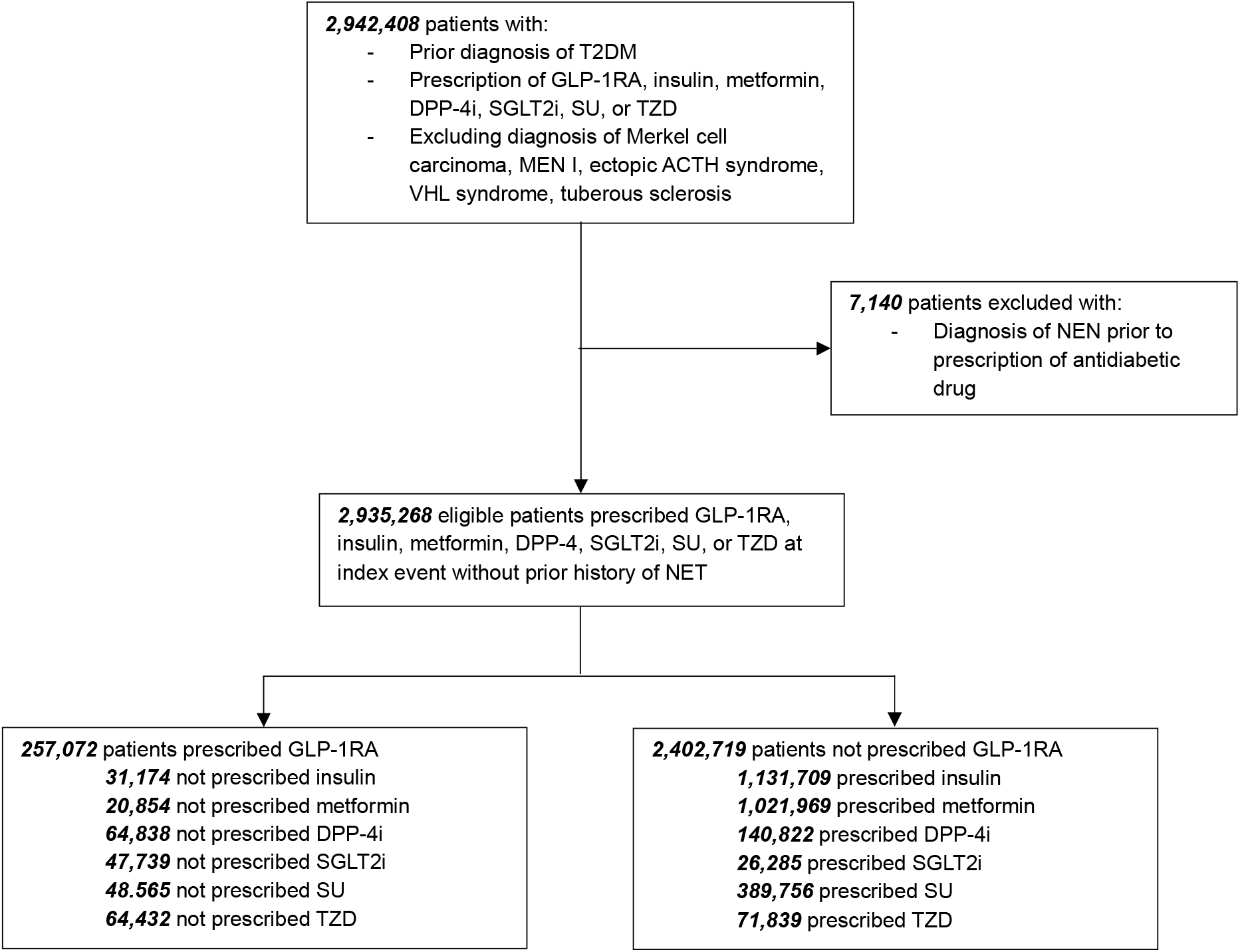
Study Cohort Selection Criteria. Diagram demonstrating cohort selection and exclusion criteria with associated population numbers. The sum of each medication group may not be equal to the total number of eligible patients as patients may be prescribed multiple antihyperglycemic agents.

